# Should rapid antigen tests be government funded in Australia? An economic evaluation

**DOI:** 10.1101/2022.01.03.22268709

**Authors:** Jonathan Karnon, Hossein Afzali, Billie Bonevski

## Abstract

**Objective:** Easy and equitable access to testing is a cornerstone of the public health response to COVID-19. Currently in Australia, testing using Polymerase Chain Reaction (PCR) tests for COVID-19 is free-to-the-user, but the public purchase their own Rapid Antigen Tests (RATs). We conduct an economic analysis of government-funded RATs in Australia.

**Design:** An interactive decision tree model was developed to compare one policy in which government-funded RATs are free-to-the-user, and one in which individuals purchase their own RATs. The decision tree represents RAT and PCR testing pathways for a cohort of individuals without COVID-19-like symptoms, to estimate the likelihood of COVID-19 positive individuals isolating prior to developing symptoms and the associated costs of testing, from a government perspective.

**Data sources:** Test costs and detection rates were informed by published studies, other input parameter values are unobservable and uncertain, for which a range of scenario analyses are presented.

**Data synthesis:** Assuming 10% prevalence of COVID-19 in a cohort of 10,000 individuals who would use government-funded RATs, the model estimates an additional 464 individuals would isolate early at a cost to the government of around $52,000. Scenario analyses indicate that the incremental cost per additional COVID-19 positive individual isolating with no symptoms remains at a few hundred dollars at 5% prevalence, rising to $2,052 at 1% prevalence.

**Conclusions:** Based on the presented decision tree model, even only minor reductions in COVID-19 transmission rates due to early isolation would justify the additional costs associated with a policy of government-funded RATs.

## Introduction

The novel coronavirus SARS-CoV-2, now known as COVID-19, was declared by the World Health Organisation (WHO) a global pandemic in 2020 and continues to spread throughout the world with new more transmissible variants. As of today, over 289 million confirmed cases have been reported globally with more than 5.4 million deaths.^1^ The pandemic has also had a huge impact on the Australian economy - a cost of $311bn was reported in the 2021-22 federal budget.

Surveillance is a key component of the public health response to COVID-19 as recommended by the WHO.^2^ Since the start of the pandemic, testing and contact tracing have been the primary measures used to interrupt the spread of COVID-19.^3^ One of the benefits of population wide active testing for cases is that it allows countries to rapidly identify new cases, isolate affected individuals and their close contacts and slow further transmission of the disease.

Australia’s response has included the use of primary care and large community-based testing hubs to implement free-to-the-user Polymerase Chain Reaction (PCR) tests for COVID-19. PCR tests are conducted by healthcare workers who take a swab from the back of the nose and throat. The swab is then sent to a pathology laboratory for testing and individuals receive their results via a mobile phone text message in approximately 24 hours, although it can take days to receive the result. It is estimated that the PCR testing system has cost the Australian government $3.7 billion to deliver to date.^4^ The presentation of the highly transmissible Omicron variant in Australia, coinciding with a relaxation of public health restrictions and the social “festive season” of Christmas and New Years Eve has led to a surge in the numbers of COVID-19 cases in Australia, swamping the PCR testing system. Rapid Antigen Tests (RATs) are an alternative test for COVID-19, which could supplement the PCR testing system.

In November 2021, the Australian Therapeutic Goods Administration (TGA) approved the use of RATs which individuals can use to self-swab at home with results available within 15-20 minutes. PCR testing remains the gold standard in terms of accuracy, however the Cochrane review of RATs reported a mean true positive rate (sensitivity) of 72% for symptomatic COVID-19 cases and 58% for people with COVID-19 but without symptoms.^5^ In Australia, unlike PCR testing, RATs are not free and are available for purchase at supermarkets and pharmacies at a cost to the user of between $10-20 per test. There are concerns that some people are unable to afford RATs, particularly when multiple tests are needed, and some public health experts and social service organisations are calling for them to be made free to encourage greater uptake. The cost is also a potential cause of inequities in COVID-19 testing and isolation.

RATs are a recent introduction to the Australian COVID-19 public health response, however they have been used in other countries for a while. In countries like Singapore, the UK, France and Germany RATs are free to the user. In Singapore, the Ministry of Health (MOH) commenced mailing six RAT kits through SingPost to households in September 2021. In addition, the Ministry of Education (MOE) and Early Childhood Development Agency (ECDA) also distributed three RAT kits to all pupils and staff in ECDA-licensed pre-schools, MOE kindergartens, early intervention centres, primary schools and the primary or junior sections of special education schools. The MOH said the distribution of RAT kits form part of a broader strategy, which includes vending machines, to step up testing efforts in Singapore.^6^ In the UK, the Medicines and Healthcare products Regulatory Agency (MHRA) has granted NHS Test & Trace an exceptional use authorisation to use certain RATs as self-tests to detect infection in people who do not have any COVID-19 symptoms and who may not otherwise have been tested.^7^ In many cases, individuals apply for RATs online and the NHS mails the tests to households. However, in January 2022, the Australian federal government ruled out providing free-to-the-user RATs tests. Some state governments have announced that they may provide free RATs in their states.

In this paper, we conduct an economic analysis to determine whether federal government-funded RATs allowing the early detection and self-isolation of people with COVID-19, are a cost-effective strategy for the Australian government to consider.

## Methods

Figure 1 presents a decision tree model that describes the testing pathways for a cohort of individuals who do not have COVID-19-like symptoms to estimate the isolation status of the COVID-19 positive members of the cohort. The primary outcome is ‘early isolation’, defined as COVID-19 positive individuals who isolate prior to the development of symptoms. The cohort is defined as individuals who would use RATs if the tests were to be funded by the government. The model estimates the expected testing costs and isolation statuses of individuals using a RAT on a single calendar day.

**Figure 1.**
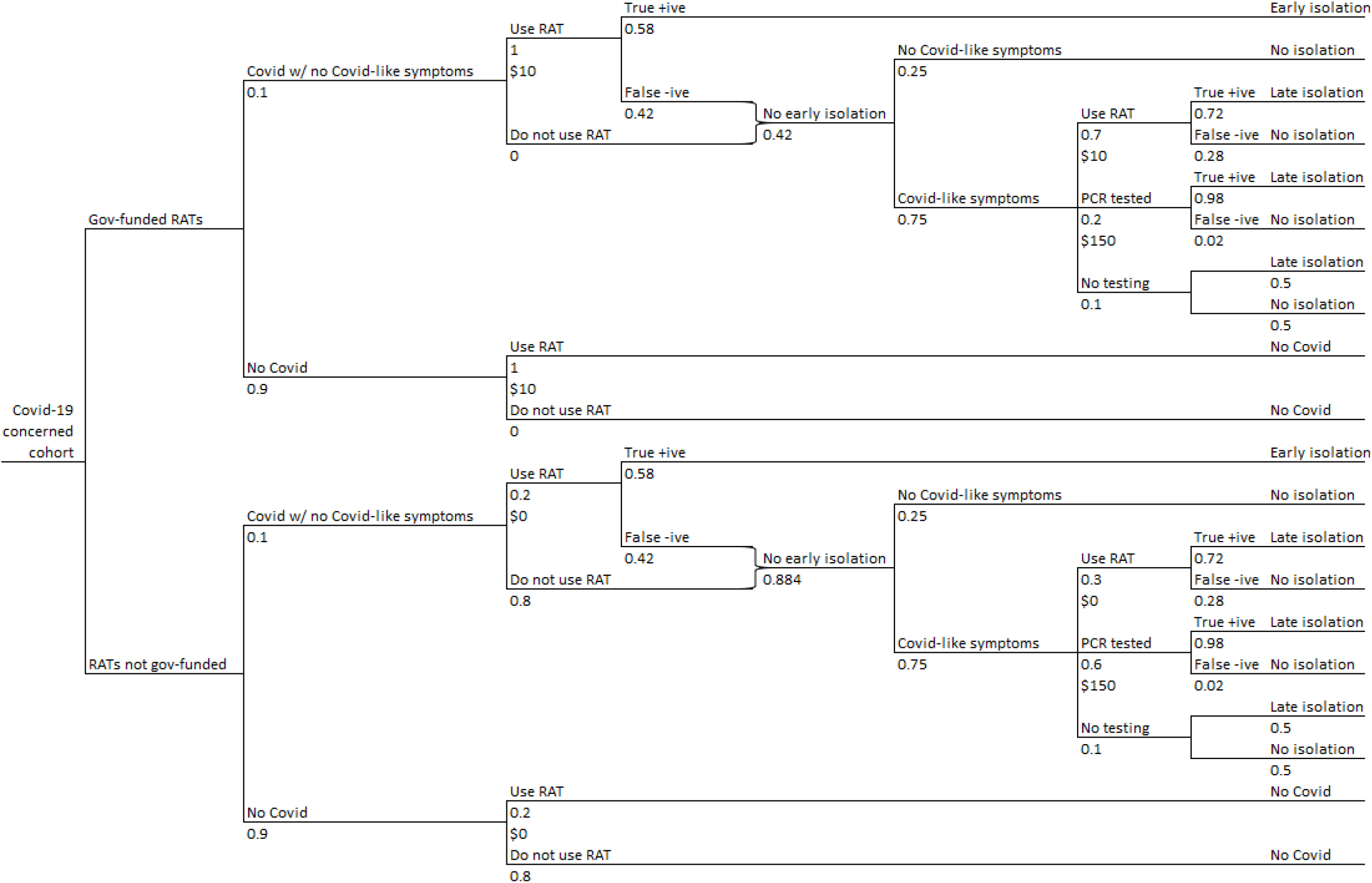
Decision tree structure.

The tree describes two policy options. The top half of the tree represents a policy in which government-funded RATs are free-to-the-user, the bottom half represents a policy under which individuals purchase their own RATs.

The next two sets of branches in the tree describe key parameters for the economic evaluation: the proportions of the defined cohort who are COVID-19 positive and negative, and the proportions of individuals who would take a RAT test with no COVID-19-like symptoms.

The RAT is not a perfect test and so the tree represents the proportion of individuals with COVID-19 who receive true positive and false negative test results. Those receiving a true positive test result are assumed to isolate early. Individuals who are not tested or who receive a false negative result may go on to experience symptoms, at which point they may be tested using a RAT or a PCR test, and depending on the test results start isolating, or not. Individuals experiencing symptoms may choose not to be tested, noting that a proportion of these individuals may still isolate.

The tree also represents the costs of RAT and PCR tests from a government perspective. All PCR testing costs are incurred by the government, whilst RAT costs are only incurred under a policy of government-funded RATs.

Table 1 presents the full set of input parameters for the model, and a set of parameter values that are used to illustrate the use of the model. Readers can access the model online at googledocs to test the effects of using alternative input parameter values.

**Table 1.** Model input parameters and illustrative parameter values.

| **Input parameters** | **Parameter values** |
| --- | --- |
| COVID-19 cases | 0.1 |
| Develop symptoms | 0.75 |
| **Use RAT \| No COVID-like symptoms** |  |
| Government-funded RATs | 1.0 |
| RATs not government-funded | 0.2 |
| **Use RAT \| COVID-like symptoms** |  |
| Government-funded RATs | 0.7 |
| RATs not government-funded | 0.3 |
| **Use PCR \| COVID-like symptoms** |  |
| Government-funded RATs | 0.2 |
| RATs not government-funded | 0.6 |
| **Isolate \| COVID symptoms & not tested** | 0.5 |
| **Test characteristics** |  |
| RAT True +ive (sensitivity) \| no symptoms | 0.58 |
| RAT True +ive (sensitivity) \| symptoms | 0.72 |
| PCR True +ive (sensitivity) | 0.98 |
| **Resource use and costs** |  |
| RAT unit cost to the government | \$10 |
| PCR test unit cost to the government | \$150 |
| Number RATs test used per day | 1 |

In the illustrative model, we assume that 10% of the ‘COVID-19 concerned cohort with no COVID-19-like symptoms’ are COVID-19 positive and that 75% will develop symptoms.

We assume the whole cohort will take a RAT test with no COVID-19-like symptoms under a policy of government-funded RATs, compared to 20% when RATs are not government funded. On developing symptoms, we assume more individuals will be tested using a PCR test if RATs are not government funded. The proportion of the cohort developing symptoms who do not get tested is assumed to be the same (10%) under both policies, as is the proportion of not tested individuals who choose to isolate.

Table 1 also describes the costs and estimated accuracy of the RAT and PCR tests, applying a lower accuracy of the RAT for individuals with COVID-19 but without symptoms.

## Results

Table 2 presents the model outputs for a cohort of 10,000 ‘COVID-19 concerned individuals’ who would use government-funded RATs for the presented illustrative set of input parameter values. The model estimates an additional 464 individuals would isolate early, prior to the development of symptoms under a policy of government-funded RATs, at a cost to the government of around $52,000. From these values, we can estimate that for every additional $112 ($51,985 divided by 464) spent by the government due to the funding of RATs, an additional individual with COVID-19 and no symptoms is expected to isolate.

**Table 2.**
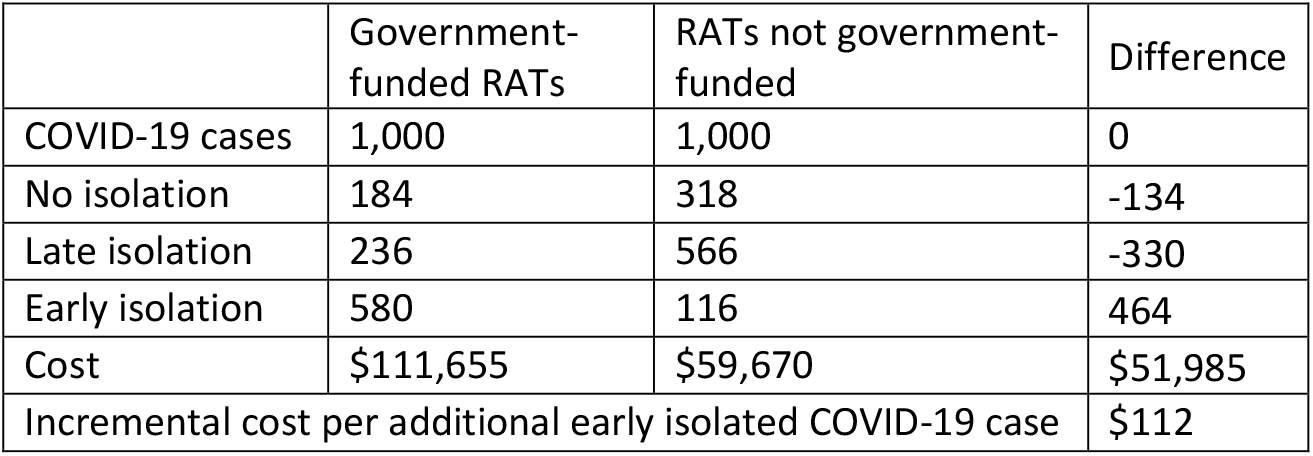
Model outputs for illustrative input parameter values.

Table 3 presents a set of scenario analyses to illustrate the effect of using alternative values for key input parameters. Decreasing the percentage of the cohort with COVID-19 to generate the largest incremental cost per additional early isolated COVID-19 case, to $328 assuming 5% prevalence, and to $2,052 assuming 1% prevalence. The other scenarios did not have a significant effect on the incremental cost per additional early isolated COVID-19 case, whilst increasing prevalence to 20% or reducing the unit cost per RAT to $5 resulted in trivial incremental costs per early isolated COVID-19 cases.

**Table 3.** Scenario analyses.

| Input parameters (base case value) | Parameter values | Additional early isolated cases | Additional Costs | ICER |
| --- | --- | --- | --- | --- |
| COVID-19 cases (0.1) | 0.01 | 46 | \$95,199 | \$2,052 |
| | 0.05 | 232 | \$75,993 | \$328 |
| | 0.2 | 928 | \$3,970 | \$4 |
| Use RAT w/ no symptoms if not gov-funded (0.2) | 0.3 | 406 | \$55,900 | \$138 |
| | 0.4 | 348 | \$59,815 | \$172 |
| RAT True +ive (sens) \| no symptoms (0.58) | 0.48 | 384 | \$53,410 | \$139 |
| | 0.68 | 544 | \$50,560 | \$93 |
| RAT cost to the government (\$10) | \$5 | 464 | \$883 | \$2 |
| | \$15 | 464 | \$103,088 | \$222 |

RAT – Rapid antigen test; ICER – incremental cost-effectiveness ratio (incremental cost per additional early isolated COVID case)

## Discussion

This paper has reported the findings from a simple cost-effectiveness analysis of government-funded RATs for the early detection of COVID-19. The presented decision tree model facilitates scenario analyses around expected testing and isolation pathways with and without government-funded RATs. Key input parameters include the prevalence of COVID-19 in individuals using RATs, the unit cost of the tests and the mean number of tests used per person tested. The prevalence of COVID-19 in individuals using RATs is unobservable, but the range of presented scenario analyses indicate that the incremental cost to the government per additional COVID-19 positive individual isolating with no symptoms would, at most, be a few hundred dollars.

The expected benefits of early isolation are difficult to quantify, but modelling by the Centre for the Mathematical Modelling of Infectious Diseases COVID modelling group provides an indication of the benefits. This group estimated that in Australia the reproductive number increased from around 1 to 1.5 over the course of December 2021, and daily COVID cases increased from a few hundred to over 30,000. This illustrates the importance of the reproductive number and the magnitude of the potential effects of increasing early isolation of people with COVID.^8,9^ Even a minor reduction in the reproductive number is expected to have significant effect on the number of daily cases.

Containing the spread of COVID-19 is important for many reasons, including the avoidance of short- and long-term health effects, reducing burden on the health system and increasing availability of essential workers. The economic benefits of accurate early identification of COVID-19 cases are clear, with earlier isolation and lower spread, businesses are likely to experience fewer staff shortages that are currently seen with staff self-isolating due to uncertainties.

All of these outcomes impose significant costs to people and society. We could make a conservative assumption that each early isolated case prevents at least one new COVID case. The broad range of costs associated with one COVID case are likely to be far higher than the estimated costs to the government per additional early isolated person with COVID.

Early isolation is an accepted strategy for containing the spread of COVID-19. In countries like Singapore and Germany, population testing and efficient vaccination roll-outs have become the key strategies of containing COVID-19 from this point forward. In addition, Mina et al (2021) argue that RAT based testing is a preferable public health measure for “test to release” because their technology allows the detection of cases that are currently infectious, as opposed to PCR based testing which results in 50-70% of positives being post-infectious, leading to unnecessarily longer quarantine times.^10^

A key concern with a user-pays model of rapid antigen self-testing is the inequities it potentially accentuates. Globally, COVID-19 is impacting negatively on vulnerable population groups,^11^ and use of expensive user-pays RATs for population testing is likely to widen the disparities. There are reports emerging in Australia of charitable organisations forced to purchase RAT kits on behalf of socioeconomically disadvantaged consumers. Thus, policy decisions regarding the distribution of RATs in Australia should also be considered through an equity lens.

Overuse of RATs by people without an underlying risk of COVID-19 would reduce the cost-effectiveness of a policy of government-funded RATs, but the unpleasant nature of the testing process should limit such overuse. In the UK, people are encouraged to do a RAT if they have been in close contact with someone with COVID-19, but also before they mix with people in crowded indoor places or visit someone at higher risk of getting seriously ill from COVID-19. Little evidence on the proportions of RAT users with COVID-19 is available. Hoarding is another risk, as with toilet paper in the early days of the pandemic and so it would be important to ensure confidence in the supply and distribution of RATs.

This paper has presented a simple cost-effectiveness analysis of government-funded RATs in Australia. The decision tree only estimated effects in terms of additional early isolated COVID-19 cases and key model input parameters are unobservable, in particular, the prevalence of COVID-19 in the cohort who would use government-funded RATs. However, our interpretation of the presented scenario analyses indicates that an implementation strategy that promotes use by people with an underlying risk of COVID-19 would be a cost-effective government policy.

## Data Availability

All data produced in the present work are contained in the manuscript.

https://tinyurl.com/mrcnuese

